# Changes in dispensing of medicines proposed for re-purposing in the first year of the COVID-19 pandemic in Australia

**DOI:** 10.1101/2021.09.26.21264150

**Authors:** Andrea L Schaffer, David Henry, Helga Zoega, Julian H Elliott, Sallie-Anne Pearson

**Author notes:** **Corresponding author:** Dr Andrea L Schaffer.

## Abstract

**Objective:** We quantified changes in dispensing of common medicines proposed for “re-purposing” due to their perceived benefits as therapeutic or preventive for COVID-19 in Australia, a country with relatively low COVID-19 incidence in the first year of the pandemic.

**Methods:** We performed an interrupted time series analysis and cross-sectional study using nationwide dispensing claims data (January 2017-November 2020). We focused on six subsidised medicines proposed for re-purposing: hydroxychloroquine, azithromycin, ivermectin, colchicine, corticosteroids, and calcitriol (Vitamin D analogue). We quantified changes in monthly dispensing and initiation trends during COVID-19 (March-November 2020) using autoregressive integrated moving average models (ARIMA) and compared characteristics of initiators in 2020 and 2019.

**Results:** In March 2020, we observed a 99% (95%CI 96%-103%) increase in hydroxychloroquine dispensing (of which approximately 22% attributable to new use), and a 199% increase (95%CI 184%-213%) in initiation, with a shift towards prescribing by general practitioners (42% in 2020 vs 25% in 2019) rather than specialists. These increases subsided following regulatory restrictions on prescribing to relevant specialties. There was a small but sustained increase in ivermectin dispensing over multiple months, with a 80% (95%CI 42%-118%) increase in initiation in May 2020 following its first identification as potentially disease-modifying in April. Other than increases in March related to stockpiling, we observed no increases in initiation of calcitriol or colchicine during COVID-19. Dispensing of corticosteroids and azithromycin remained lower than expected in April through November 2020.

**Conclusions:** While most increases in dispensing observed early on during COVID-19 were temporary and appear to be related to stockpiling among existing users, we did observed increases in initiation of hydroxychloroquine and ivermectin and a shift in prescribing patterns which may be related to media hype around these medicines. A quick response by regulators can help limit inappropriate repurposing to lessen the impact on medicine supply and patient harms.

## INTRODUCTION

Since COVID-19 was first recognised, several medicines have been proposed for ‘re-purposing’ in the belief they may be effective in treating or preventing COVID-19, the disease caused by SARS-CoV-2.[1,2] Large, well-conducted trials have confirmed potential benefits for dexamethasone, cytokine inhibitors and remdesivir.[3–6] However, other medicines (e.g. hydroxychloroquine, ivermectin, Vitamin D) have not been shown to be effective yet misinformation about their benefits has been widespread. There are reports of medicines being prescribed extensively despite a lack of robust efficacy or safety data.[7] For instance, in the United States (US) the number of new prescriptions for the antimalarial and anti-rheumatic medicine hydroxychloroquine was over 7-fold higher in March 2020 than in the previous year, with a shift in the characteristics of prescribers and people using the medicine.[8]

Worldwide, the success of measures to reduce the spread of the virus has varied. In 2020, Australia had a relatively low incidence of COVID-19 compared with other countries, with a cumulative incidence of approximately 28,000 cases (109 per 100,000 population) and 908 deaths as of December 2020.[9] Studies of changing medicine use patterns early on in the pandemic focussed on high COVID-19 incidence settings.[8,10] The relatively low Australian infection rate means that substantial increases in use of medicines believed to be of benefit in COVID-19 during this time period are unlikely to be due to management of confirmed COVID-19; they more likely reflect stockpiling over concerns about supply shortages by people already using the medicine, or new use among people who believe in their preventive effect.

The early days of the COVID-19 pandemic were associated with great uncertainty; it is important to understand how large-scale health crises impact on medicine use so that policy-makers can act quickly to promote quality use of medicines and prevent harms. Given the ever-changing evidence, and some misinformation, around effective care of people with COVID-19, the Australian government established the National COVID-19 Clinical Evidence Taskforce, a multi-disciplinary collaboration between researchers and clinicians. Its role is to undertake continuous evidence surveillance and develop ‘living’ evidence-based guidelines, including recommendations for use of prescribed medicines in treating or preventing COVID-19.[11]

Our primary objective was to examine changes in use and initiation of widely available medicines in Australia, several of which featured in media coverage because of their claimed benefits in preventing or treating COVID-19. We examined changes in monthly dispensing and treatment initiation in March through November 2020 and compared the characteristics of people initiating these medicines before and during COVID-19.

## METHODS

### Context

Australia maintains a publicly funded, universal healthcare system entitling all citizens and eligible residents to subsidised prescribed medicines through the Pharmaceutical Benefits Scheme (PBS). Medicines dispensed by community pharmacies are known as the general schedule, or Section 85 (S85). In 2020, Australia had a low overall COVID-19 incidence but experienced two notable spikes in cases in March and July.[9] We selected March 2020 as the interruption point as this coincided with a nationwide emergency response plan and marked the start of the initial nationwide lockdown[9].

### Medicines of interest

We focused on six prescribed medicines available on the PBS general schedule (S85), that were proposed for re-purposing for prevention or treatment of COVID-19, most of which were the subject of extensive media coverage.[12–14] We used the National COVID-19 Clinical Evidence Taskforce’s Australian guidelines for the clinical care of people with COVID-19[11] as of July 2021 to guide judgment about which of several categories applied: 1) recommended for use (corticosteroids), 2) should be used only in a clinical trial (ivermectin, calcitriol [Vitamin D analogue]) or 3) should not be used for prevention or treatment of COVID-19 (hydroxychloroquine, azithromycin, colchicine) (**Table S1**). We also looked at use of hydroxychloroquine and azithromycin combined, which was reported early on as a potentially beneficial combination.[2]

### Data sources

We used two sources of PBS data. First, we used publicly-available, monthly aggregate claims for all S85 medicines dispensed to PBS-eligible persons from January 2017 to November 2020 to analyse overall changes in dispensing after March 2020.[15] While these data capture all community dispensing in Australia, they do not contain person-level characteristics. For more detailed analyses, we used person-level claims for a 10% random sample of all PBS-eligible people for the same period. These data contain information on medicines dispensed, including prescriber specialty, and the patient’s year of birth and sex. The 10% sample is a standard dataset provided by Services Australia for analytical use and is selected based on the last digit of each person’s randomly assigned unique identifier. To protect privacy, dispensing dates are offset by +/-14 days; the offset is the same within each individual. PBS dispensing data mostly reflect prescribing in general practice with a small proportion from specialists in their offices, private hospital inpatients and aged care residents. PBS claims do not capture medicines dispensed to public hospital inpatients or private dispensings (i.e., not PBS-subsidised where the consumer pays the entire cost out-of-pocket).

### Statistical analyses

We used the aggregated PBS data to quantify changes in all S85 medicines combined and each medicine of interest from March to November 2020. We used interrupted time series analyses with autoregressive integrated moving average (ARIMA) models to estimate monthly changes from March to November 2020.[16] Details on the methodology are in the **Appendix**. To account for stockpiling, we summed the change in the number of dispensings predicted by the model over all months to estimate the total change during the COVID-19 period. We estimated 95% confidence intervals (CIs) by summing lower and upper bounds of the estimated change in each month.

Second, using person-level data for 10% of PBS-eligible people, we examined patterns of dispensing and treatment initiation for each medicine of interest. We defined initiation as the first observed dispensing after a period of 360 days without dispensing of that medicine. We performed interrupted time series analysis using ARIMA models as described above to quantify changes in the number of initiators. For medicines where we observed ≥1 month with a significant increase in initiation between March and November 2020, we compared the characteristics of people initiating (sex, age, prescriber specialty) to initiators during the same period in 2019.

In December 2019 and January 2020, Australia also experienced severe bushfires that may have impacted on prescribed medicine use.[17] Therefore, we tested inclusion of dummy variables representing this period in the modelling. As the impact was minimal, we removed the bushfire covariate from the final models.

We performed all analyses with R V4.0.2 and SAS V9.4.

### Ethics and data access approvals

This study was approved by the New South Wales Population and Health Services Research Ethics Committee (no. 2013/11/494). The Australian Government Services Australia External Request Evaluation Committee granted access for the 10% sample of PBS claims for the study (no. RMS1126).

## RESULTS

### Overall dispensing

We observed a 20.0% (95%CI 17.0% to 23.0%) increase over the predicted estimates in all S85 PBS medicines dispensed in March 2020 of 5,110,790 (95%CI 4,350,937 to 5,870,644). This was followed by several months of decreased dispensing (**Figure 1, Table S2**).

**Figure 1.**
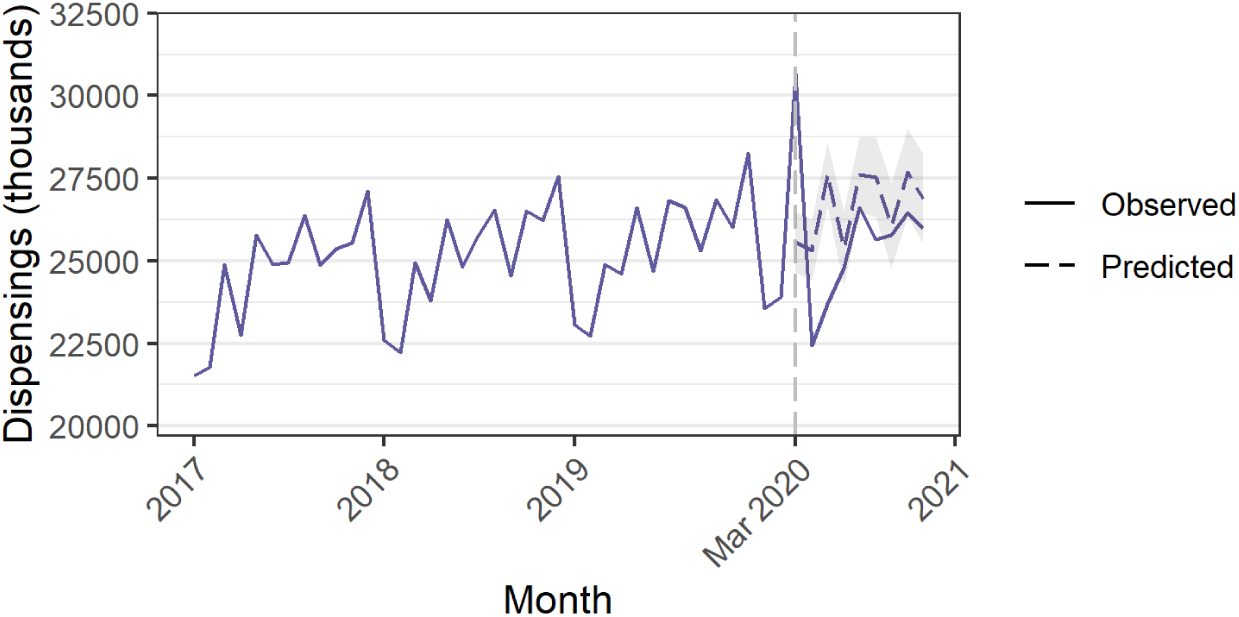
Dispensing of all medicines. Grey shaded area = 95% CI for predicted values

### Azithromycin

We did not observe an increase in azithromycin dispensing in March 2020 (Table S2). Azithromycin dispensing was lower than expected in April through November, and we observed 61,766 fewer dispensings during the COVID-19 period (95%CI -72,457 to -51,075) (**Figure 2A; Table 1)**. Similarly, in the PBS 10% sample data, we observed fewer people initiating azithromycin each month from April through November (**Figure 3A; Table S3**).

**Table 1.** Change in dispensing from March to November 2020 estimated using autoregressive integrated moving average models (ARIMA) with full aggregate s85 dispensing data

|  | Median monthly dispensings in 2019 | Total change in dispensing, Mar-Nov 2020 |  |  |  |
| --- | --- | --- | --- | --- | --- |
|  |  | N | 95% CI | % | 95% CI |
| **All dispensings** | 25 666 813 | -7 508 685 | -15 967 327 to 949 957 | -3.1% | -6.7% to 0.4% |
| Azithromycin | 22 699 | -61 766 | -72 457 to -51 075 | -28.6% | -33.5% to -23.6% |
| Hydroxychloroquine | 25 481 | 9195 | 538 to 17 852 | 3.9% | 0.2% to 7.5% |
| Ivermectin | 977 | 1923 | 344 to 3501 | 22.3% | 4.0% to 40.6% |
| Colchicine | 48 767 | 12 230 | -2510 to 26 969 | 2.7% | -0.6% to 5.9% |
| Corticosteroids | 366 886 | -767 236 | -1 107 670 to -426 801 | -22.1% | -31.9% to -12.3% |
| Calcitriol | 11 413 | 4610 | 340 to 8879 | 4.4% | 0.3% to 8.5% |

**Figure 2.**
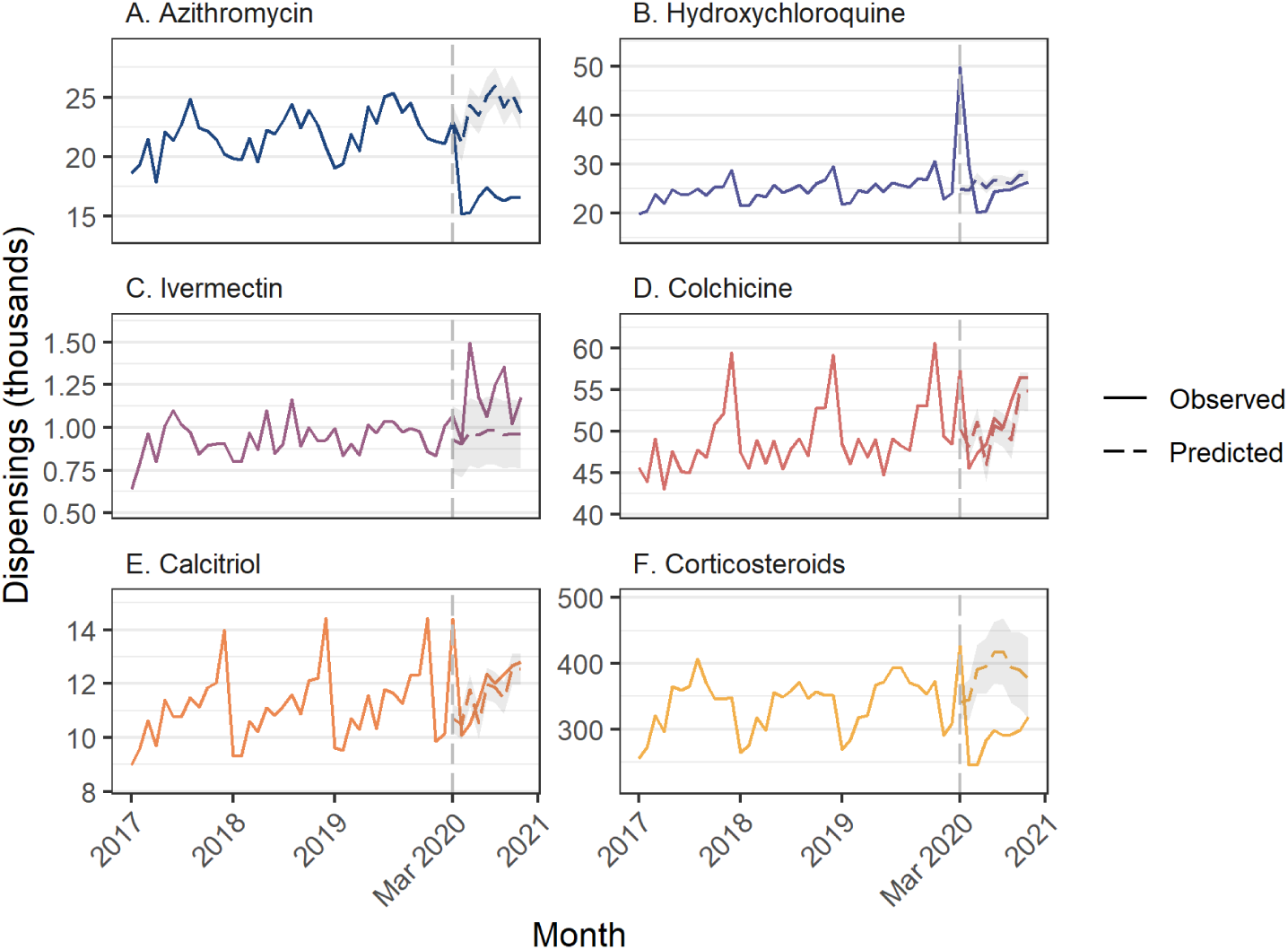
Dispensing of medicines of interest using full aggregate Section 85 dispensing data. Grey shaded area = 95% CI for predicted values.

**Figure 3.**
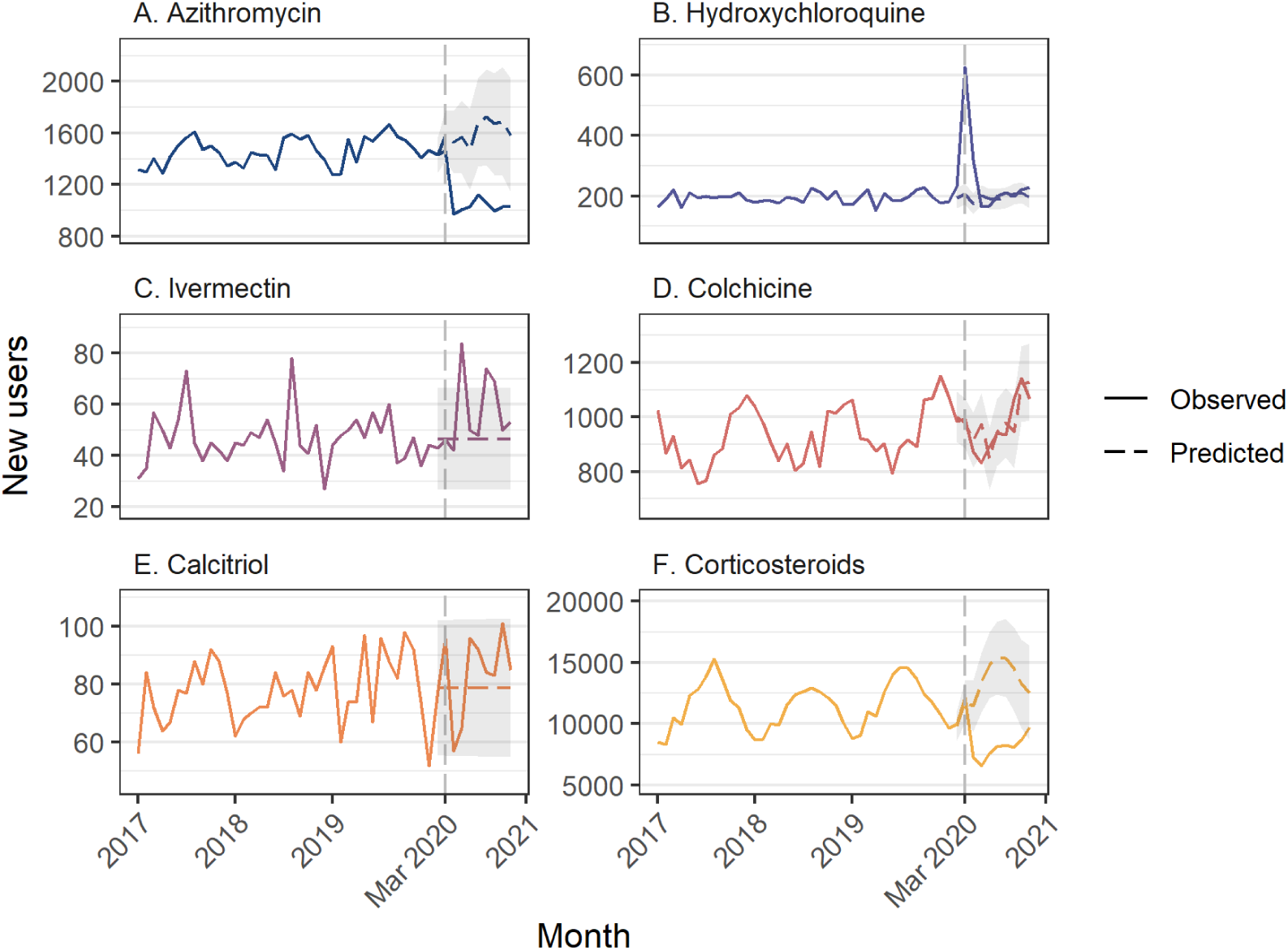
Number of new users of each medicine class of interest estimated using 10% sample of PBS dispensing claims data. New user = new dispensing without a previous dispensing in the past 360 days. Grey shaded area = 95% CI for predicted values.

### Hydroxychloroquine

We observed an increase in hydroxychloroquine dispensings of 24,799 (95%CI 23,887 to 25,711) in March 2020, representing a 99.4% increase (95%CI 95.8% to 103.1%) over the predicted value, and an increase of 4977 (95%CI 4042 to 5912) in April. (**Figure 2B, Table S2**). While hydroxychloroquine dispensings were lower than predicted in May through October, they did not offset the earlier increases; there was an estimated 9195 more hydroxychloroquine dispensings (95%CI 538 to 17,852) than predicted over the COVID-19 period (March through November) (**Table 1**).

In the 10% PBS sample, we observed a monthly median of 2438 hydroxychloroquine dispensings and 281 people initiating in 2019. In March 2020, 415 additional people initiated hydroxychloroquine (95%CI 385 to 446), a 198.6% increase (95%CI 184.1%-213.1%) (**Figure 3B, Table S3**). In the 10% PBS sample we estimated that there were an additional 1884 dispensings in March 2020 (**Table S4**), meaning that roughly 78% of the spike in dispensings in March 2020 was likely due to stockpiling among people previously treated with the medicine. Over the COVID-19 period, there were an additional 562 people with new hydroxychloroquine use from March to November (95%CI 292 to 832) (**Table 2**).

**Table 2.** Change in number of initiators from March to November 2020 estimated using autoregressive integrated moving average models (ARIMA). Estimates are based on a 10% sample of PBS-eligible people.

|  | Median monthly new users in 2019 | Total change in initiation, Mar-Nov 2020 |  |  |  |
| --- | --- | --- | --- | --- | --- |
|  |  | No. new users | 95% CI | % | 95% CI |
| Azithromycin | 1540 | -4282 | -5068 to -3496 | -30.6% | -36.2% to -24.9 % |
| Hydroxychloroquine | 196 | 562 | 292 to 832 | 31.7% | 16.4% to 46.9% |
| Ivermectin | 48 | 97 | -63 to 256 | 23.1% | -14.9% to 61.2% |
| Colchicine | 916 | -91 | -974 to 792 | -1.0% | -11.0% to 9.0% |
| Corticosteroids | 12 080 | -45 986 | -67 768 to -24 204 | -37.6% | -55.3% to -19.8% |
| Calcitriol | 85 | 54 | -135 to 242 | 7.6% | -19.1% to 34.2% |

We observed a significant increase in initiation of hydroxychloroquine therapy in March and April 2020, so we compared the characteristics of people initiating the medicine in these months with the same period in 2019. Of people with new hydroxychloroquine use in 2020, 30.1% were male compared with 23.4% during the same months in 2019 (**Table 3**). We observed a substantial increase in the number of new hydroxychloroquine dispensings written by GPs; in 2020, 41.8% (n=533) were written by GPs compared with 24.6% (n=129) in 2019. Conversely, the proportion of prescriptions written by rheumatologists was 45.3% (n=238) in 2019 and 25.8% (n=567) in 2020. Combined use of hydroxychloroquine and azithromycin was rare, with only 16 initiators of hydroxychloroquine also prescribed azithromycin in the same month during COVID-19.

**Table 3.**
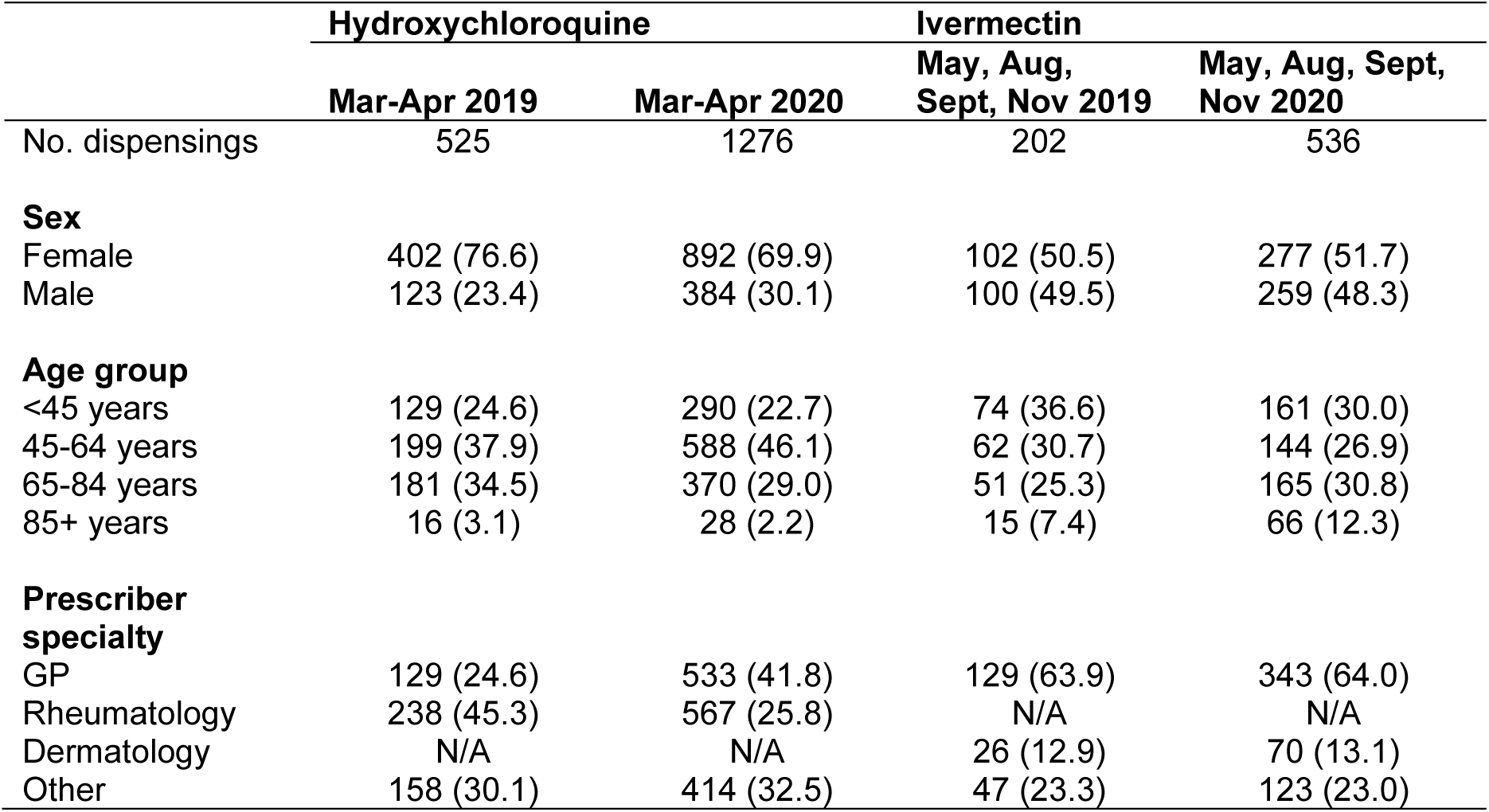
People initiating hydroxychloroquine and ivermectin in months in 2020 where there was a greater than expected number of new users, compared with new users in 2019. Estimates are based on a 10% sample of PBS dispensing claims data.

|  | Hydroxychloroquine |  | Ivermectin |  |
| --- | --- | --- | --- | --- |
|  | Mar-Apr 2019 | Mar-Apr 2020 | May, Aug, Sept, Nov 2019 | May, Aug, Sept, Nov 2020 |
| No. dispensings | 525 | 1276 | 202 | 536 |
| **Sex** |  |  |  |  |
| Female | 402 (76.6) | 892 (69.9) | 102 (50.5) | 277 (51.7) |
| Male | 123 (23.4) | 384 (30.1) | 100 (49.5) | 259 (48.3) |
| **Age group** |  |  |  |  |
| <45 years | 129 (24.6) | 290 (22.7) | 74 (36.6) | 161 (30.0) |
| 45-64 years | 199 (37.9) | 588 (46.1) | 62 (30.7) | 144 (26.9) |
| 65-84 years | 181 (34.5) | 370 (29.0) | 51 (25.3) | 165 (30.8) |
| 85+ years | 16 (3.1) | 28 (2.2) | 15 (7.4) | 66 (12.3) |
| **Prescriber specialty** |  |  |  |  |
| GP | 129 (24.6) | 533 (41.8) | 129 (63.9) | 343 (64.0) |
| Rheumatology | 238 (45.3) | 567 (25.8) | N/A | N/A |
| Dermatology | N/A | N/A | 26 (12.9) | 70 (13.1) |
| Other | 158 (30.1) | 414 (32.5) | 47 (23.3) | 123 (23.0) |

### Ivermectin

We did not observe any increase in ivermectin dispensing in March 2020, but there was greater than expected ivermectin dispensing in May, June, August, September, and November 2020, with the largest increase in May 2020 (520, 95%CI 344 to 696, a 53.3% increase compared with predicted) (**Figure 2C, Table S2**). Overall, there were 1923 more dispensings than expected during COVID-19 (95% CI 344 to 3501) (**Table 1**).

In the PBS 10% sample, there was a monthly median of 90 ivermectin dispensings and 52 people initiating therapy in 2019. In May 2020 an additional 37 people initiated treatment (95%CI 20 to 55), a 80.4% increase (95%CI 42.3%-118.4%); however, there was no overall increase in the number of people initiating new treatment over the COVID-19 period (**Figure 3C, Table 2, Table S3**).

As we observed a significant increase in new use of ivermectin in May, August, September, and November, we compared the characteristics of initiators in this period in 2020 with 2019. While the number of people initiating therapy was greater, there were few differences in their characteristics. In both 2019 and 2020 approximately 64% of new dispensings were prescribed by GPs.

### Colchicine

We observed an increase in colchicine dispensings in March of 7106 (95%CI 5814 to 8397) (**Figure 2D, Table S1**). Smaller increases were also observed in June and September; however, there was no overall change in dispensings over the COVID-19 period (12,230, 95%CI -2510 to 26,969) (**Table 1**). We did not observe any months with an increase in people initiating of colchicine treatment (**Table S2**).

### Calcitriol (vitamin D analogue)

We observed increases in calcitriol dispensings in March, June, and September 2020, with the greatest increase in March (3698, 95%CI 3256 to 4140, a 34.5% increase) (**Figure 2E, Table S2**). In the 10% PBS sample data there was no change in the number of people initiating therapy in any month (**Figure 2E, Table S3**).

### Corticosteroids

We observed fewer corticosteroid dispensings in all months from April through November (**Figure 2F, Table S2**). Overall, there were 767,736 fewer dispensings than predicted (95%CI -1,107,670 to -426,801). In the 10% PBS sample, we observed a similar pattern with a decrease in initiation in every month from April through October (**Figure 3F, Table S3**), with no increase in any month.

## DISCUSSION

During the COVID-19 epidemic in Australia as elsewhere, claims have been made about the unproven benefits of several common medicines. Australia provides a unique context for studying changes in use of medicines proposed for repurposing for treatment of prevention of COVID-19 at the start of the pandemic due to the initial low incidence rate, meaning that use of these medicines to treat people with confirmed cases of COVID-19 would be very rare. In this study we found significant increases in dispensing of some of these medicines, but these trends were partly explained by stockpiling by people already on therapy, likely in anticipation of supply shortages and less access to medical practitioners.

As has been documented elsewhere[18,19] dispensing of all PBS-listed medicines rose by approximately 20% in March 2020, followed by a decrease lasting several months, attributed to stockpiling by people receiving treatment for a broad range of disorders. Stockpiling has been observed internationally as well.[20,21] While to our knowledge there have been no reports of people with legitimate needs for these medicines being unable to access them,[22] nor increases in adverse events, stockpiling does put pressure on medicine supply and may exacerbate shortages. This is particularly true during crises such as the COVID-19 pandemic which led to disruptions of the global supply chain through lockdowns, understaffing and travel bans.[23] While there are many causes for shortages, local regulatory agencies and policymakers can play a role in mitigating their short-term impacts.[24,25] This may include identifying which medicines are most vulnerable to shortages, restricting the conditions under which they may be prescribed to limit waste and overuse, and acting quickly in response to changing circumstances.[26,27]

Hydroxychloroquine was one of the earliest PBS-listed medicine subjected to speculation in the media as a potential anti-viral treatment.[7] Hydroxychloroquine is used mainly as chronic treatment for autoimmune disorders and has been shown to be ineffective in treating or preventing COVID-19; the Australian National Clinical Evidence Taskforce recommends against its use.[28,29] We observed a large spike in hydroxychloroquine dispensing early in the pandemic, with roughly 70% driven by stockpiling by people already on treatment, some of whom may have been concerned by shortages. However, the greatest relative increase was related to new use, and more likely to have been prescribed by a GP rather than a specialist which is consistent with findings from the US, where a 10.5-fold increase in new prescriptions by primary care physicians was observed.[8] On March 24, due to concerns over off-label prescribing of hydroxychloroquine the Therapeutic Goods Administration (TGA) which regulates medicines in Australia, limited who could initiate therapy to relevant specialties[30] and following an update by the Taskforce that hydroxychloroquine was “not recommended” on April 30, the TGA further increased restrictions on prescribing.[31] We did not have data on prescribing indication but another Australian study found that only half of people newly prescribed hydroxychloroquine by GPs during COVID-19 had a relevant condition in their medical history.[32]

Azithromycin, another treatment that received a ‘do not use’ recommendation by the Taskforce has been widely promoted as COVID-19 treatment as a sole therapy or in combination with hydroxychloroquine. There was no increase in dispensing at the start of the pandemic, but we observed a sustained fall through the remainder of the study period, a pattern also observed with corticosteroids; this has been noted for many antibiotics and is related to a reduction in respiratory infections during COVID-19 restrictions.[33,34] We found few cases of co-dispensing of azithromycin and hydroxychloroquine among people initiating hydroxychloroquine, which is also consistent with previously reported general practice data.[32]

In contrast to hydroxychloroquine, the spikes in use of ivermectin, a widely promoted therapy whose use is currently not recommended outside trials by the National COVID-19 Clinical Evidence Taskforce nor the World Health Organisation,[11,35] occurred later in the pandemic and were more distributed across time. Its role as potentially disease-modifying in COVID-19 was not publicised until April 2020.[36,37] We did not observe any ivermectin stockpiling by people already on therapy, as it is typically taken to treat scabies as a one-off treatment. However, the changes seen during the COVID-19 pandemic in Australia—an increase of around 1900 dispensings, on an annual background of 12,000—suggest a modest uptake in the belief that it can treat or prevent COVID-19. Ivermectin has continued to attract media attention with several recent clinical trials plagued by problems such as errors or fabrication.[38] Lastly, while vitamin D has been promoted as COVID-19 treatment it is not recommended by the Taskforce for use outside clinical trials. Our data were limited to dispensing of calcitriol, which did not exhibit much change in dispensing during the COVID-19 epidemic.

### Strengths and limitations

We had complete capture of medicine dispensing for the whole Australian population, and person-level data on a 10% sub-sample. However, we do not have data on dispensing of private prescriptions, meaning we have likely underestimated the impact on use of some medicines. We also did not have information on the indication for prescribing and cannot determine whether use was actually off-label or related to COVID-19. However, given the very low incidence of COVID-19 during the study period, and that these data primarily represent community dispensing, it is likely that only a tiny minority of increases in use of these medicines were for treatment of COVID-19, but more likely represent a response to media attention and/or stockpiling, as we observed changes in pattern of use of some medicines (such as hydroxychloroquine) not consistent with typical use. Lastly, disruptions to medicine use during COVID-19 likely have multiple causes, including lockdown measures, changed interaction with the healthcare system, reduced circulation of respiratory and gastrointestinal infections, and fear of not being able to access medicines, and these cannot reliably be disentangled.

## Conclusions

We demonstrated temporary changes in dispensing of commonly used medicines that were proposed for re-purposing for the treatment and prevention of COVID-19 early in the pandemic, including a large short-lived increase in hydroxychloroquine dispensing, most of which may be due to anticipatory stockpiling, and a later smaller but longer-lasting increase in ivermectin dispensing. Balanced and informed communication of the changing evidence, including up-to-date and reliable access to evidence-informed advice is necessary to minimise any negative health impacts related to re-purposing of medicines. When similar situations arise, a quick response by regulators can help limit inappropriate re-purposing, to avoid supply shortages and potential harms.

## Data Availability

Aggregate data underlying the findings of this study are provided in a supplementary file (S3). Section 85 PBS claims data are publicly available at https://www.pbs.gov.au/info/statistics/dos-and-dop/dos-and-dop. The 10% sample of person-level PBS claims data were used under licensed from the Australian Gvoernment Services Australia. Access to person-level data by other individuals or authorities is not permitted without the express permission of the approving human research ethics committees and data custodians.

## Acknowledgements

This research is supported by the National Health and Medical Research Council (NHMRC) Centre of Research Excellence in Medicines Intelligence (#1196900). Dr Schaffer is supported by a NHMRC Early Career Fellowship (#1158763). A/Prof Zoega is supported by a UNSW Scientia Fellowship. We thank the Australian Government Services Australia for providing the data. Thank you to Prof Andrew Wilson for his input.

## References

1. Bartoszko JJ, Siemieniuk RAC, Kum E, Qasim A, Zeraatkar D, Ge L, et al. Prophylaxis against covid-19: living systematic review and network meta-analysis. BMJ. 2021;373:n949. doi:10.1136/bmj.n949

2. Sanders JM, Monogue ML, Jodlowski TZ, Cutrell JB. Pharmacologic Treatments for Coronavirus Disease 2019 (COVID-19): A Review. JAMA. 2020;323: 1824–1836. doi:10.1001/jama.2020.6019

3. RECOVERY Collaborative Group, Horby P, Lim WS, Emberson JR, Mafham M, Bell JL, et al. Dexamethasone in Hospitalized Patients with Covid-19. N Engl J Med. 2021;384: 693–704. doi:10.1056/NEJMoa2021436

4. Tomazini BM, Maia IS, Cavalcanti AB, Berwanger O, Rosa RG, Veiga VC, et al. Effect of Dexamethasone on Days Alive and Ventilator-Free in Patients With Moderate or Severe Acute Respiratory Distress Syndrome and COVID-19: The CoDEX Randomized Clinical Trial. JAMA. 2020;324: 1307–1316. doi:10.1001/jama.2020.17021

5. Salama C, Han J, Yau L, Reiss WG, Kramer B, Neidhart JD, et al. Tocilizumab in Patients Hospitalized with Covid-19 Pneumonia. N Engl J Med. 2021;384: 20–30. doi:10.1056/NEJMoa2030340

6. Spinner CD, Gottlieb RL, Criner GJ, Arribas López JR, Cattelan AM, Soriano Viladomiu A, et al. Effect of Remdesivir vs Standard Care on Clinical Status at 11 Days in Patients With Moderate COVID-19: A Randomized Clinical Trial. JAMA. 2020;324: 1048–1057. doi:10.1001/jama.2020.16349

7. Gabler E, Keller MH. Prescriptions Surged as Trump Praised Drugs in Coronavirus Fight. The New York Times. 25 Apr 2020. Available: https://www.nytimes.com/2020/04/25/us/coronavirus-trump-chloroquine-hydroxychloroquine.html. Accessed 27 Apr 2020.

8. Bull-Otterson L, Gray EB, Budnitz DS, Strosnider HM, Schieber LZ, Courtney J, et al. Hydroxychloroquine and Chloroquine Prescribing Patterns by Provider Specialty Following Initial Reports of Potential Benefit for COVID-19 Treatment - United States, January-June 2020. MMWR Morb Mortal Wkly Rep. 2020;69: 1210–1215. doi:10.15585/mmwr.mm6935a4

9. COVID-19 National Incident Room Surveillance Team. COVID-19 Australia: Epidemiology Report 31: Reporting period ending 6 December 2020. Communicable Diseases Intelligence. 2020;44. doi:10.33321/cdi.2020.44.92

10. Geller AI, Lovegrove MC, Lind JN, Datta SD, Budnitz DS. Assessment of Outpatient Dispensing of Products Proposed for Treatment or Prevention of COVID-19 by US Retail Pharmacies During the Pandemic. JAMA Intern Med. 2021;181: 869–872. doi:10.1001/jamainternmed.2021.0299

11. National COVID-19 Clinical Evidence Taskforce. National COVID-19 Clinical Evidence Taskforce. 2021 [cited 2 Mar 2021]. Available: https://covid19evidence.net.au/

12. McGinn C. Coronavirus Australia: Ivermectin, Anti-parasitic drug kills COVID-19 in lab. news.com.au. 3 Apr 2020. Available: https://www.news.com.au/lifestyle/health/health-problems/coronavirus-australia-ivermectin-antiparasitic-drug-kills-covid19-in-lab/news-story/615c435e56aefc4b704f4fd890bd4c2c. Accessed 7 Jul 2021.

13. Davey M. Decades-old drug in two Australian trials related to Covid-19 but experts urge caution. The Guardian. 24 Mar 2020. Available: http://www.theguardian.com/australia-news/2020/mar/24/decades-old-drug-in-two-australian-trials-related-to-covid-19-but-experts-urge-caution. Accessed 7 Jul 2021.

14. Kekatos M. Gout drug may improve survival odds for COVID-19 patients, study says. In: Daily Mail Online [Internet]. 24 Jun 2020 [cited 7 Jul 2021]. Available: https://www.dailymail.co.uk/health/article-8455395/Could-old-gout-drug-help-treat-coronavirus-Study-suggests-colchicine-fight-inflammation.html

15. Australian Government Department of Health. PBS and RPBS Section 85 Date of Supply Data. Available: https://www.pbs.gov.au/info/statistics/dos-and-dop/dos-and-dop

16. Schaffer AL, Dobbins TA, Pearson S-A. Interrupted time series analysis using autoregressive integrated moving average (ARIMA) models: a guide for evaluating large-scale health interventions. BMC Medical Research Methodology. 2021;21: 58. doi:10.1186/s12874-021-01235-8

17. Borchers Arriagada N, Palmer AJ, Bowman DM, Morgan GG, Jalaludin BB, Johnston FH. Unprecedented smoke-related health burden associated with the 2019–20 bushfires in eastern Australia. The Medical Journal of Australia. 2020;213: 282–283. doi:10.5694/mja2.50545

18. Australian Institute of Health and Welfare. Impacts of COVID-19 on Medicare Benefits Scheme and Pharmaceutical Benefits Scheme service use. In: Australian Government Australian Institute of Health and Welfare [Internet]. 2020 [cited 2 Mar 2021]. Available: https://www.aihw.gov.au/reports/health-care-quality-performance/covid-impacts-on-mbs-and-pbs/contents/summary

19. Mian M, Sreedharan S, Giles S. Increased dispensing of prescription medications in Australia early in the COVID-19 pandemic. The Medical Journal of Australia. 2021;214: 428–429. doi:10.5694/mja2.51029

20. Karlsson P, Nakitanda AO, Löfling L, Cesta CE. Patterns of prescription dispensation and over-the-counter medication sales in Sweden during the COVID-19 pandemic. PLOS ONE. 2021;16: e0253944. doi:10.1371/journal.pone.0253944

21. Enners S, Gradl G, Kieble M, Böhm M, Laufs U, Schulz M. Utilization of drugs with reports on potential efficacy or harm on COVID-19 before, during, and after the first pandemic wave. Pharmacoepidemiology and Drug Safety. n/a. doi:10.1002/pds.5324

22. Tang M, Daniels B, Aslam M, Schaffer A, Pearson S-A. Changes in systemic cancer therapy in Australia during the COVID-19 pandemic: a population-based study. The Lancet Regional Health – Western Pacific. 2021;14. doi:10.1016/j.lanwpc.2021.100226

23. Socal MP, Sharfstein JM, Greene JA. The Pandemic and the Supply Chain: Gaps in Pharmaceutical Production and Distribution. Am J Public Health. 2021;111: 635–639. doi:10.2105/AJPH.2020.306138

24. Burry LD, Barletta JF, Williamson D, Kanji S, Maves RC, Dichter J, et al. It Takes a Village…: Contending With Drug Shortages During Disasters. Chest. 2020;158: 2414–2424. doi:10.1016/j.chest.2020.08.015

25. Kuo S, Ou H-T, Wang CJ. Managing medication supply chains: Lessons learned from Taiwan during the COVID-19 pandemic and preparedness planning for the future. Journal of the American Pharmacists Association. 2021;61: e12–e15. doi:10.1016/j.japh.2020.08.029

26. Badreldin HA, Atallah B. Global drug shortages due to COVID-19: Impact on patient care and mitigation strategies. Research in Social and Administrative Pharmacy. 2021;17: 1946–1949. doi:10.1016/j.sapharm.2020.05.017

27. Qato DM, Ozenberger K, Olfson M. Prevalence of Prescription Medications With Depression as a Potential Adverse Effect Among Adults in the United States. JAMA. 2018;319: 2289–2298. doi:10.1001/jama.2018.6741

28. Boulware DR, Pullen MF, Bangdiwala AS, Pastick KA, Lofgren SM, Okafor EC, et al. A Randomized Trial of Hydroxychloroquine as Postexposure Prophylaxis for Covid-19. N Engl J Med. 2020;383: 517–525. doi:10.1056/NEJMoa2016638

29. Abella BS, Jolkovsky EL, Biney BT, Uspal JE, Hyman MC, Frank I, et al. Efficacy and Safety of Hydroxychloroquine vs Placebo for Pre-exposure SARS-CoV-2 Prophylaxis Among Health Care Workers: A Randomized Clinical Trial. JAMA Intern Med. 2021;181: 195–202. doi:10.1001/jamainternmed.2020.6319

30. Australian Government Therapeutic Goods Administration. New restrictions on prescribing hydroxychloroquine for COVID-19. In: Therapeutic Goods Administration (TGA) [Internet]. Australian Government Department of Health; 24 Mar 2020 [cited 16 Apr 2021]. Available: https://www.tga.gov.au/alert/new-restrictions-prescribing-hydroxychloroquine-covid-19

31. Australian Government Department of Health. Revised hydroxychloroquine PBS listings for the treatment of autoimmune disorders and malaria from 1 May 2020. Australian Government Department of Health; Available: https://www.pbs.gov.au/info/news/2020/05/revised-hydroxychloroquine-pbs-listings-for-the-treatment

32. Hydroxychloroquine: did COVID-19 change GP prescribing? In: NPS MedicineWise [Internet]. [cited 26 Mar 2021]. Available: https://www.nps.org.au/news/hydroxychloroquine-did-covid-19-change-gp-prescribing

33. Sullivan SG, Carlson S, Cheng AC, Chilver MB, Dwyer DE, Irwin M, et al. Where has all the influenza gone? The impact of COVID-19 on the circulation of influenza and other respiratory viruses, Australia, March to September 2020. Eurosurveillance. 2020;25: 2001847. doi:10.2807/1560-7917.ES.2020.25.47.2001847

34. Gillies MB, Burgner DP, Ivancic L, Nassar N, Miller JE, Sullivan SG, et al. Changes in antibiotic prescribing following COVID-19 restrictions: Lessons for post-pandemic antibiotic stewardship. British Journal of Clinical Pharmacology. n/a. doi:10.1111/bcp.15000

35. World Health Organization. Therapeutics and COVID-19: living guideline. WHO; 2021 Sep. Available: https://www.who.int/publications-detail-redirect/WHO-2019-nCoV-therapeutics-2021.3

36. Monash Biomedicine Discovery Institute. Lab experiments show anti-parasitic drug, Ivermectin, eliminates SARS-CoV-2 in cells in 48 hours. 3 Apr 2020 [cited 24 Mar 2021]. Available: https://www.monash.edu/discovery-institute/news-and-events/news/2020-articles/Lab-experiments-show-anti-parasitic-drug,-Ivermectin,-eliminates-SARS-CoV-2-in-cells-in-48-hours

37. Caly L, Druce JD, Catton MG, Jans DA, Wagstaff KM. The FDA-approved drug ivermectin inhibits the replication of SARS-CoV-2 in vitro. Antiviral Research. 2020;178: 104787. doi:10.1016/j.antiviral.2020.104787

38. Schraer R, Goodman J. Ivermectin: How false science created a Covid miracle drug. BBC News. 6 Oct 2021. Available: https://www.bbc.com/news/health-58170809. Accessed 7 Oct 2021.

