## Supplementary material for "Changes in dispensing of medicines proposed for re-purposing in the first year of the COVID-19 pandemic in Australia": Table S1

### SUPPLEMENTARY TABLES

**Table S1:** List of pharmaceutical treatments reviewed by the National COVID-19 taskforce as of July 2021 and their availability in Australia

| Medicine name | National COVID-19 Clinical Evidence Taskforce recommendation | Availability in Australia* |
|---|---|---|
| Azithromycin | Do not use | PBS-subsidised (S85) |
| Hydroxychloroquine | Do not use | PBS-subsidised (S85) |
| Ivermectin | Do not use outside clinical trials | PBS-subsidised (S85) |
| Colchicine | Do not use | PBS-subsidised (S85) |
| Corticosteroids | Use | PBS-subsidised (S85) |
| Calcitriol | Do not use outside clinical trials | PBS-subsidised (S85); other vitamin D analogues such as calcifediol are available over-the-counter |

PBS = Pharmaceutical Benefits Scheme; S85 = Section 85 (Section 85 is the general schedule and refers to items dispensed by community pharmacies)

**Table S2.** Monthly change in dispensing in 2020 compared with predicted values estimated using ARIMA models with full aggregate Section 85 dispensing data

| Month in 2020 | Azithromycin n (95% CI) | Hydroxychloroquine n (95% CI) | Ivermectin n (95% CI) | Colchicine n (95% CI) |
|---|---|---|---|---|
| Mar | 69 (-1033 to 1171) | 24799 (23887 to 25711) | 142 (-31 to 316) | 7106 (5814 to 8397) |
| Apr | -5867 (-7042 to -4692) | 4977 (4042 to 5912) | 22 (-154 to 198) | -2640 (-3936 to -1344) |
| May | -8883 (-10142 to -7624) | -7032 (-8025 to -6038) | 520 (344 to 696) | -3888 (-5232 to -2545) |
| Jun | -6748 (-7922 to -5575) | -4736 (-5742 to -3731) | 221 (47 to 396) | 2457 (799 to 4114) |
| Jul | -7533 (-8796 to -6269) | -2382 (-3379 to -1386) | 80 (-97 to 257) | 880 (-774 to 2533) |
| Aug | -9229 (-10404 to -8055) | -1898 (-2871 to -926) | 263 (86 to 439) | 231 (-1523 to 1984) |
| Sep | -7804 (-8995 to -6613) | -1287 (-2240 to -335) | 399 (224 to 574) | 4695 (2812 to 6578) |
| Oct | -8640 (-9808 to -7472) | -2048 (-3004 to -1091) | 55 (-120 to 231) | 1743 (-127 to 3613) |
| Nov | -7132 (-8316 to -5948) | -1197 (-2130 to -265) | 220 (45 to 395) | 1647 (-344 to 3638) |

| Month in 2020 | Corticosteroids n (95% CI) | Calcitriol n (95% CI) | All medicines n (95% CI) |
|---|---|---|---|
| Mar | 87195 (66643 to 107747) | 3698 (3256 to 4140) | 5110790 (4350937 to 5870644) |
| Apr | -98233 (-124238 to -72229) | -418 (-888 to 52) | -2865463 (-3623925 to -2107001) |
| May | -145302 (-176357 to -114246) | -1283 (-1756 to -810) | -3924526 (-4705203 to -3143850) |
| Jun | -112895 (-148176 to -77614) | 830 (355 to 1305) | -582967 (-1531222 to 365288) |
| Jul | -117706 (-156781 to -78631) | 367 (-110 to 844) | -1002106 (-1951755 to -52458) |
| Aug | -127310 (-169837 to -84783) | 148 (-332 to 628) | -1881239 (-2871030 to -891448) |
| Sep | -102474 (-148194 to -56754) | 866 (384 to 1348) | -268893 (-1339225 to 801440) |
| Oct | -90971 (-139675 to -42267) | 139 (-345 to 623) | -1217241 (-2295849 to -138633) |
| Nov | -59540 (-111056 to -8025) | 262 (-225 to 749) | -877041 (-2000055 to 245974) |

ARIMA = autoregressive integrated moving average

**Table S3.** Monthly change in initiators in 2020 compared with predicted values estimated using ARIMA models with a 10% sample of PBS data where the dispensing date is offset by +/- 14 days

| Month in 2020 | Azithromycin n (95% CI) | Hydroxychloroquine n (95% CI) | Ivermectin n (95% CI) | Colchicine n (95% CI) |
|---|---|---|---|---|
| Mar | -61 (-148 to 27) | 415 (385 to 446) | -1 (-18 to 17) | 14 (-64 to 91) |
| Apr | -510 (-597 to -423) | 150 (119 to 182) | -5 (-22 to 13) | -42 (-126 to 42) |
| May | -500 (-588 to -411) | -37 (-67 to -8) | 37 (20 to 55) | -142 (-235 to -50) |
| Jun | -380 (-471 to -290) | -21 (-51 to 8) | 3 (-14 to 21) | 46 (-47 to 138) |
| Jul | -503 (-589 to -416) | 11 (-18 to 41) | 1 (-16 to 19) | -1 (-101 to 100) |
| Aug | -603 (-690 to -516) | 17 (-13 to 46) | 27 (10 to 45) | -44 (-148 to 60) |
| Sep | -620 (-706 to -533) | -12 (-42 to 19) | 22 (5 to 40) | 121 (15 to 227) |
| Oct | -610 (-697 to -523) | 6 (-25 to 37) | 3 (-14 to 21) | 21 (-91 to 132) |
| Nov | -496 (-583 to -410) | 33 (3 to 62) | 6 (-11 to 24) | -63 (-178 to 52) |

| Month in 2020 | Corticosteroids n (95% CI) | Calcitriol n (95% CI) |
|---|---|---|
| Mar | 358 (-1077 to 1793) | 18 (-13 to 49) |
| Apr | -4157 (-5914 to -2400) | -35 (-69 to -1) |
| May | -6778 (-8807 to -4749) | -11 (-47 to 25) |
| Jun | -7229 (-9498 to -4960) | 26 (-12 to 64) |
| Jul | -7217 (-9702 to -4732) | 18 (-22 to 58) |
| Aug | -7121 (-9805 to -4437) | -1 (-43 to 41) |
| Sep | -6411 (-9281 to -3541) | -5 (-49 to 39) |
| Oct | -4611 (-7655 to -1567) | 46 (0 to 92) |
| Nov | -2820 (-6028 to 388) | 21 (-26 to 68) |

ARIMA = autoregressive integrated moving average

**Table S4.** Monthly change in dispensing in 2020 compared with predicted values estimated using ARIMA models with 10% sample of PBS data where the dispensing date is offset by +/- 14 days

| Month in 2020 | Azithromycin n (95% CI) | Hydroxychloroquine n (95% CI) | Ivermectin n (95% CI) | Colchicine n (95% CI) |
|---|---|---|---|---|
| Mar | 89 (-53 to 231) | 316 (222 to 411) | -7 (-34 to 20) | 217 (96 to 337) |
| Apr | -28 (-231 to 174) | 1884 (1785 to 1984) | -7 (-34 to 21) | 479 (358 to 599) |
| May | -616 (-862 to -369) | 750 (646 to 855) | -12 (-40 to 15) | -28 (-149 to 92) |
| Jun | -702 (-988 to -416) | -391 (-500 to -281) | 48 (21 to 76) | -165 (-312 to -18) |
| Jul | -536 (-857 to -215) | -353 (-467 to -239) | 23 (-5 to 50) | 47 (-106 to 201) |
| Aug | -794 (-1142 to -446) | -184 (-302 to -65) | 1 (-27 to 29) | 188 (35 to 341) |
| Sep | -788 (-1164 to -412) | -115 (-238 to 8) | 22 (-6 to 50) | -62 (-232 to 108) |
| Oct | -859 (-1260 to -457) | -109 (-236 to 18) | 64 (36 to 91) | 358 (189 to 527) |
| Nov | -931 (-1356 to -505) | -91 (-222 to 40) | 23 (-5 to 50) | 63 (-119 to 244) |

| Month in 2020 | Corticosteroids n (95% CI) | Calcitriol n (95% CI) |
|---|---|---|
| Mar | 1787 (175 to 3399) | 154 (53 to 256) |
| Apr | 5686 (3406 to 7966) | 263 (159 to 368) |
| May | -6983 (-9776 to -4190) | 31 (-76 to 139) |
| Jun | -12010 (-15235 to -8785) | -68 (-178 to 41) |
| Jul | -11372 (-14977 to -7767) | 91 (-22 to 203) |
| Aug | -10177 (-14126 to -6228) | 22 (-93 to 138) |
| Sep | -11139 (-15405 to -6873) | 36 (-81 to 154) |
| Oct | -9876 (-14436 to -5316) | 41 (-79 to 162) |
| Nov | -7491 (-12328 to -2654) | 111 (-11 to 234) |

ARIMA = autoregressive integrated moving average
