## Appendix for "Changes in dispensing of medicines proposed for re-purposing in the first year of the COVID-19 pandemic in Australia"

### Autoregressive integrated moving average (ARIMA) modelling

For a detailed description of ARIMA modelling, please see Schaffer *et al.*[1] We provide a brief overview below.

Many time series exhibit seasonality and/or autocorrelation, which violate the assumptions of ordinary least squares (OLS) regression if not appropriately controlled. Thus, for this project we chose to use ARIMA modelling based on our previous experience with Pharmaceutical Benefit Scheme (PBS) data. While ARIMA is used to model continuous data, and our data consists of counts (i.e. number of dispensings or new users), a Poisson distribution can be approximated by a Normal distribution when the expected counts ($\lambda$) are large and the distribution is not bounded by zero.

In contrast to linear regression which includes time as a predictor, ARIMA models have a single dependent variable ($Y_t$) that is a function of past values of $Y$ and the error term ($\varepsilon_t$) only. An ARIMA model combines an autoregressive (AR) component, a differencing component (I, also called "integration"), and a moving-average (MA) component to induce stationarity. A stationarity time series has constant mean, variance, and covariance, and thus is easier to predict. An ARIMA model is specified by ($p$,$d$,$q$), where $p$ is the number of autoregressive terms, $d$ is the order of differencing, and $q$ is the order of the moving-average. If data are seasonal, a seasonal ARIMA model can be used which is specified by $(p,d,q)\text{x}(P,D,Q)_S$, where $D$ is the seasonal order of differencing, and $P$ and $Q$ are the AR and MA components of the seasonal model, and $S$ is the seasonality (e.g. 12 for monthly date).

To identify the most appropriate orders for $p$ and $q$ that gave the best fitting model, we used the function *auto.arima()* in the forecast package in R.[2] Based on preliminary visualisation, for series that exhibited trends we prespecified a first difference ($d$=1). For series that exhibited seasonality, we prespecified a first seasonal difference ($D$=1). The values for $p$, $q$, $P$, and $Q$ were chosen by *auto.arima()* to minimise the AIC and BIC. Having the lowest AIC and/or BIC does not guarantee a good fitting model; therefore, we verified that the model selected by the algorithm met the regression modelling assumptions, specifically that the residuals were normally distributed, with constant variance (no heteroscedasticity), and no residual autocorrelation. To check the first two assumptions, we visualised the plots of residuals against time and residuals against fitted values, as well as normal quantile plots. To check for residual autocorrelation, we examined the autocorrelation and partial autocorrelation plots of the residuals and used the Ljung-Box test for white noise up to lag 12. The null hypothesis of the Ljung-Box test is that the data are independently distributed and that any observed correlation is a result of randomness rather than serial correlation. If any of these assumptions were violated, we chose different values for *p*, *q*, *P*, and/or *Q* and rechecked the model fit. We estimated the *p*, *d*, *q, P, D, Q* orders separately for each model being estimated. The ARIMA model specifications for each model are listed in Table 1, and an example of the R code used is in Box 1.

To estimate the change in dispensing compared with the counterfactual (i.e. predicted dispensing had previous trends continued), we included dummy variables representing each month during the COVID-19 period (March through November 2020 for analyses using the Section 85 data). As the date of dispensing is offset in the person-level data, we also included a dummy variable for February 2020. To determine the percentage change, we calculated the difference between the observed and predicted values. To visualise the difference between the observed and predicted values had the trends prior to COVID-19 continued, we truncated the time series at February 2020 and estimated the predicted values and their 95% confidence intervals using the *forecast()* function in the forecast package and the ARIMA specification identified above.

**References**


1. Schaffer AL, Dobbins TA, Pearson S-A. Interrupted time series analysis using autoregressive integrated moving average (ARIMA) models: a guide for evaluating large-scale health interventions. BMC Med Res Methodol. 2021 Mar 22;21(1):58.

2. Hyndman RJ, Khandakar Y. Automatic Time Series Forecasting: The forecast Package for R. J Stat Softw. 2008 Jul 29;27(1):1–22.
